# Biopsychosocial Phenogroups in Individuals with Coronary Artery Disease and their Associated Cardiovascular Mortality Risks

**DOI:** 10.1101/2025.09.28.25336851

**Authors:** Jeffery Osei, Baffour Otchere, Louis Li, Kara Suvada, Luis C. Correia, J. Douglas Bremner, Arshed A. Quyyumi, Yan V. Sun, Viola Vaccarino, Amit J. Shah

## Abstract

**Background:** Patients with stable coronary artery disease (CAD) represent a clinically heterogeneous group, with varying psychosocial, metabolic, and cardiovascular profiles that may differentially influence prognosis. We sought to identify distinct clinical phenogroups among patients with stable CAD and to evaluate their associations with cardiovascular disease (CVD)-specific and all-cause mortality.

**Methods:** We pooled data from 949 participants with stable CAD enrolled in two related studies. To identify distinct clinical phenogroups, we applied a model-based clustering approach using markers of autonomic dysfunction, psychosocial stress burden, myocardial injury and obstructive burden, left ventricular (LV) systolic dysfunction, and blood pressure. All variables were assessed at study enrollment. Hazard ratios (HRs) were estimated to examine the associations between the derived phenotypes and both CVD-specific and all-cause mortality.

**Results:** The mean (SD) age of participants was 60 (±10) years; 34% were women and 41% were Black. We identified four phenogroups with differing sociodemographic and clinical profiles. Compared with phenogroup 2 (low-risk factor burden group), phenogroups 1 (cardiac autonomic dysfunction group) and 3 (ischemic cardiomyopathy group) had a 2 to 10-fold higher risk of CVD-specific and all-cause mortality over a median 5 years of follow-up. These associations remained strong and statistically significant in cluster 3 even after adjustment for demographic factors. Phenogroup 4 (high psychosocial burden group) showed a more modest but consistent 1.4 to 2.3-fold higher risk of CVD-specific and all-cause mortality compared with phenogroup 2, though this did not reach statistical significance.

**Conclusions:** Our study identified novel phenogroups of CAD patients with clinically meaningful differences in risk for CVD-specific and all-cause mortality. Although more studies are warranted for this first-in-kind study, these results support a holistic model of CAD evaluation, with the promise of improving outcomes through targeted, personalized therapies that include behavioral interventions.

## Introduction

Individuals with stable coronary artery disease (CAD) represent a clinically heterogeneous group, with varying psychosocial, metabolic, and cardiovascular profiles that may differentially influence prognosis. Recent advances in unsupervised machine learning have enabled sophisticated phenotyping approaches that can identify clinically meaningful patient subgroups, or phenogroups, based on complex, multidimensional data. Phenogroups allow us to broaden our focus on clinical pathophysiology and risk, and instead of viewing each risk factor in isolation. By understanding patients as part of a phenogroup, we can obtain a more holistic view of the patient, account for risk factor clustering, and better individualize our care.^1, 2^ This is particularly important as we consider behavioral therapies such as cardiac rehabilitation that can be customized to focus on stress, physical activity, and nutrition, for example.^3^

Measures of psychological and physiological stress may improve our understanding of CAD pathophysiology and help us understand which patients may be most likely to benefit from certain behavioral interventions.^4, 5^ Unfortunately, relatively few studies have incorporated this into their phenomapping algorithms, given the challenges of collecting psychosocial data in large cohorts.^6^ Nonetheless, certain measures like patient reported outcomes and resting heart rate variability are increasingly available using free tools and consumer wearable devices.^7^ Because of previous research showing that measures of psychological stress have strong prognostic value, it is likely that groups in which stress mechanisms are dominant will have increased risk of mortality.^4^ Furthermore, by examining them in a cluster analysis alongside other risk factors, we can better understand them within a broader context and gain a better conceptual understanding of patients, even when full psychological profiles are not available.

In this study, we sought to derive novel phenogroups in patients with CAD that participated in two studies with similar protocols that included detailed psychological characterization. To enhance potential clinical translation, we limited our dataset to traditional CVD predictors, autonomic measures, and psychological self-reported stress that could be obtained in most clinical settings. We hypothesized that psychosocial and autonomic factors would play a key role in the formation of these groups, that they would vary by age and sex, and that they would be differentially associated with CVD mortality.

## Methods

### Study population and design

Our study sample was drawn from 2 related studies, the MIMS2 (Myocardial Infarction and Mental Stress Study 2; n=313) and MIPS (Mental Stress Ischemia Mechanisms and Prognosis Study; n=636).^8^ Both of which recruited participants with CAD and adhered to similar protocols as previously described.^9^ While MIMS2 recruited post-MI patients within 8 months of their event, MIPS recruited from a larger pool of participants with stable CAD. The Emory University Institutional Review Board approved the research protocol for both studies, and all participants provided written informed consent during enrollment.^8^

### Data Collection

Sociodemographic factors (age, sex, race, income, employment, and current smoking), medical history (hypertension, hyperlipidemia, diabetes mellitus, obesity, heart failure, previous percutaneous transluminal coronary angioplasty, and previous coronary artery bypass graft), and list of medications (antidepressants, beta blockers, aspirin and statin) taken were assessed using standardized questionnaires and assisted by trained clinical staff. Depressive symptoms were assessed via the Beck Depression Inventory II, a 21-item self-report assessment of depressive symptoms.^10^ Current PTSD symptoms were assessed using the PTSD symptom checklist (PCL), a 17-item self-report measure of the 17 Diagnostic Statistical Manual IV symptoms of PTSD (range, 17–85).^11^ The 10-item version of the Everyday Discrimination Scale (EDS) was used to assess exposure to everyday discrimination, or everyday occurrences of unfair treatment.^12^ Sample items include being treated with less courtesy than other people, receiving poorer service than other people at restaurants or stores, etc., without reference to race, gender, or any other characteristic. The EDS has been widely used with good reliability and validity across racial/ethnic groups.^13^ Angina symptoms were assessed using the Seattle Angina Questionnaire’s (SAQ) angina frequency subscale, which measures frequency of angina and use of nitroglycerin for chest pain over the previous four weeks. The SAQ has been validated in different populations and correlates with electronic daily diary entries for angina and nitroglycerin use.^14^

CAD severity was quantified using the Gensini semiquantitative angiographic scoring system, which takes into account the potential functional significance of the coronary lesion with a multiplier for specific coronary tree locations.^15^

Each participant wore a Holter monitor (GE Marquette SEER digital system; GE Medical Systems, Waukesha, WI) while in a seated position. Heart rate variability (HRV) measures were obtained in a 5-minute window period while at rest using GE MARS 8.0.2 (2015) software. Fast Fourier transformation was used to classify HRV into 2 frequency bands: LF (0.05 to <0.15 Hz) and HF (0.15 to <0.40 Hz).

Blood was drawn while at rest for hs-Troponin-I (hs-cTnI) measurement. Samples were processed and stored at −80°C. Plasma hs-cTnI was measured using the ARCHITECT STAT Hs-cTnI assay (Abbott Laboratories, Abbott Park, Illinois), which has a limit of detection of 1.2 pg/ml and an interassay coefficient of variation of <10% at 4.7 pg/ml.

Follow-up data were collected through patient contacts, medical record review, and the Social Security Death Index. Medical records were obtained and reviewed for hospitalizations. In MIPS, participants were contacted every 6 months for the first 3 years and then at the 5-year mark. In MIMS2, participants were contacted at the 3- and 5-year anniversary from their initial visit (approximately). All events were adjudicated by cardiologists who were blinded to their baseline study outcomes. The primary endpoint was cardiovascular death, and all-cause death was the secondary endpoint.

### Model-based clustering approach

We used a data-driven, risk factor-based clustering approach, specifically, model-based clustering with parameterized finite Gaussian mixture models to identify distinct clinical phenogroups. This method assumes that the observed data arise from a heterogeneous population composed of several latent subpopulations (clusters) and utilizes the expectation-maximization (EM) algorithm for maximum-likelihood (ML).^16–18^ We evaluated 12 continuous variables measured at baseline during rest, including markers of autonomic dysfunction (LF and HF HRV, heart rate), myocardial injury and obstructive burden (hs-cTnI, Gensini score, angina frequency), left ventricular (LV) systolic dysfunction (ejection fraction), blood pressure, and psychosocial stress burden (discrimination score, Beck Depression Inventory, and the PTSD score) for the clustering algorithm. These variables were chosen based on our findings from previous research and feasibility to obtain them in clinical settings. All variables were standardized before clustering. Variables with missing data were imputed using multiple imputations by chained equations.

For model-based clustering, we used a soft assignment framework, whereby each observation was assigned a probability of belonging to each cluster. The final cluster membership for each observation was then determined by assigning each observation to the cluster with the highest posterior probability. This mixture model was then used to partition the data into clusters by assigning each observation to the cluster for which its posterior probability of membership is highest. The optimal model and number of clusters were determined using the Bayesian Information Criterion (BIC). The Mclust package (version 6.1.1) in R (version 4.5.1; The R Foundation for Statistical Computing, Vienna, Austria) was used to perform the model-based clustering algorithm.^19^

### Statistical analysis

Baseline sociodemographic and clinical characteristics were compared across clusters using analysis of variance for continuous variables and chi-square tests for categorical variables. These comparisons are valid since cluster assignment was based on posterior probabilities from the model-based clustering algorithm. Time-to-event analyses for cardiovascular and all-cause mortality were performed using cumulative incidence curves and Cox proportional hazards regression, adjusting for age, sex, and race. Hazard ratios with 95% confidence intervals were calculated using the lowest-risk cluster as reference.

## Results

The overall cohort had a mean age of 60 (±10) years; 34% were women and 41% were Black. Over a median follow-up of 5.9 years, 51 (5%) and 90 (9%) CVD-specific and all-cause mortality were recorded, respectively.

### Model selection and cluster formation

First, we examined the correlations among the variables and found no strong correlations among them (–0.5 < r < 0.5). The results of the model-based cluster analysis are shown in Figure 1. The optimal model involved a four-cluster solution with a BIC of −23736.85. This model consisted of four clusters with the VVE covariance parameter (variable volume, variable shape, and equal orientation). The average posterior probabilities for the most likely cluster classification were 0.987, 0.945, 0.926, and 0.939 for clusters 1, 2, 3, and 4, respectively. This indicates a high degree of certainty in the classification of the four clusters.

**Figure 1.**
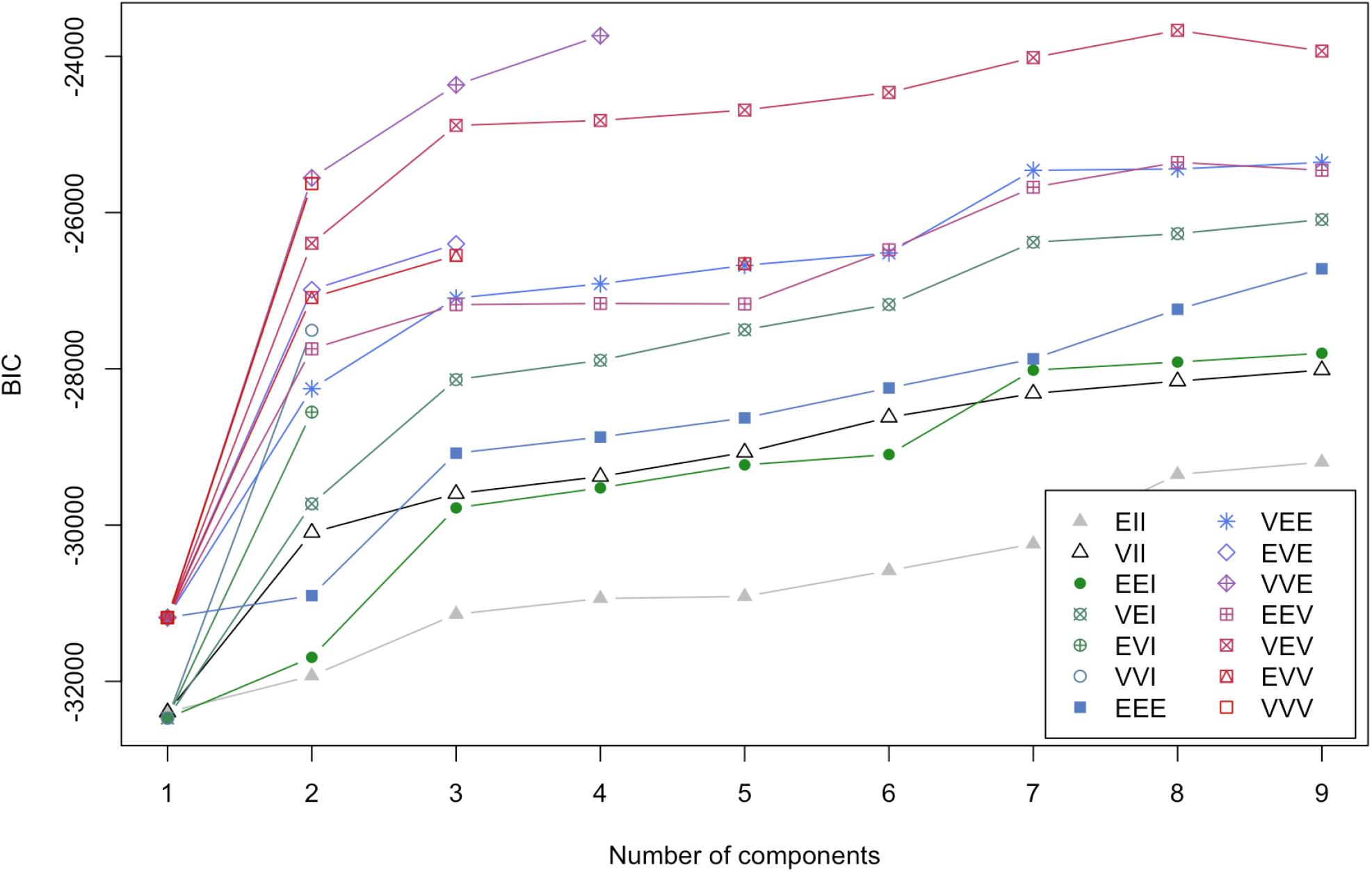
Bayesian Information Criterion (BIC) of different models and cluster indices. Unique symbol/color patterns represent the identifiers for various models used in the parameterization of the covariance matrix. Briefly, EII represents a spherical model with equal volume and shape; VII, a spherical model with variable volume but equal shape; EEI, diagonal, equal volume and shape; VEI, diagonal with variable volume and equal shape; EVI, diagonal, equal volume but variable shape; VVI, diagonal, variable volume and shape; EEE, general model, equal volume, shape and orientation; VEE, general, variable volume, equal shape and orientation; EVE, general, equal volume and orientation but variable shape; VVE, general, variable volume and shape, equal orientation; EEV, general, equal volume and shape, variable orientation; VEV, general, variable volume and orientation, equal shape; EVV, general, equal volume, variable shape and orientation; VVV, general, variable volume, shape and orientation. The optimal model uses the VVE covariance parameter, which identified four clusters and has a BIC of −23736.85.

### Phenogroup characteristics

The four phenogroups (clusters) demonstrated distinct sociodemographic and clinical characteristics (Table 1). Phenogroup 1 comprised the oldest participants and had the highest prevalence of heart failure. Phenogroup 2 included the highest proportion of White and employed participants, with the highest income levels, and the lowest prevalence of prior CABG and heart failure. Phenogroup 3 was the youngest group, with the greatest proportion of males and the highest prevalence of prior CABG. In contrast, phenogroup 4 had higher female representation, had the highest proportion of Black participants and low-income individuals (family income <$20,000/year), and the highest prevalence of antidepressant users.

**Table 1.**
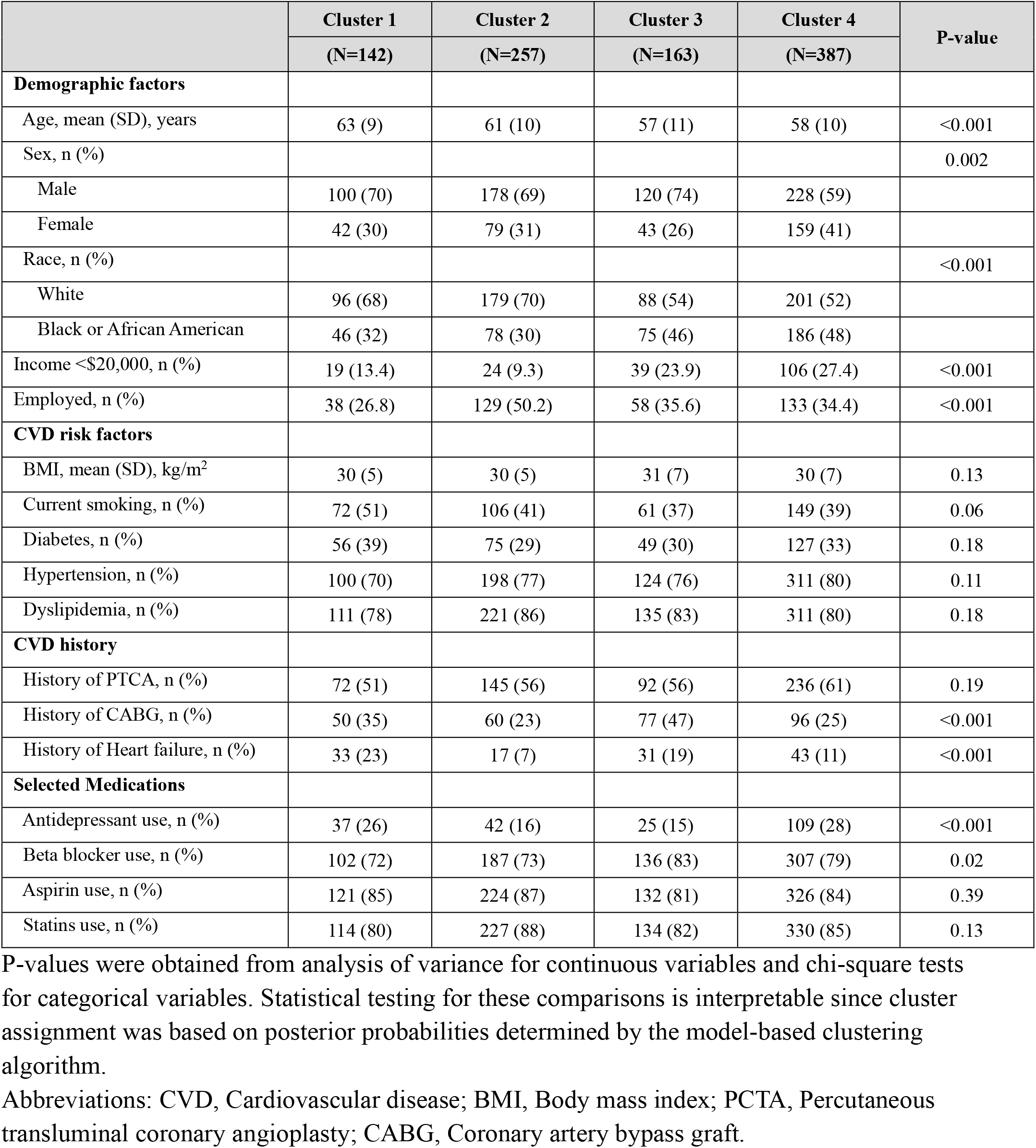
Participant characteristics.

As illustrated in Figure 2 and summarized in Table 2, the phenogroups also differed across the clinical domains. Phenogroup 1, the “cardiac autonomic dysfunction” group, was characterized by elevated levels of autonomic dysfunction and myocardial injury/obstructive burden. Phenogroup 2, the “low risk factor burden” group, consistently had the lowest levels across all five domains. Phenogroup 3, the “ischemic cardiomyopathy” group, demonstrated a substantial burden of myocardial injury and obstructive symptoms. Finally, phenogroup 4, the “high psychosocial burden” group, exhibited the highest levels of psychosocial stress, alongside low to intermediate levels of the other domains.

**Table 2.**
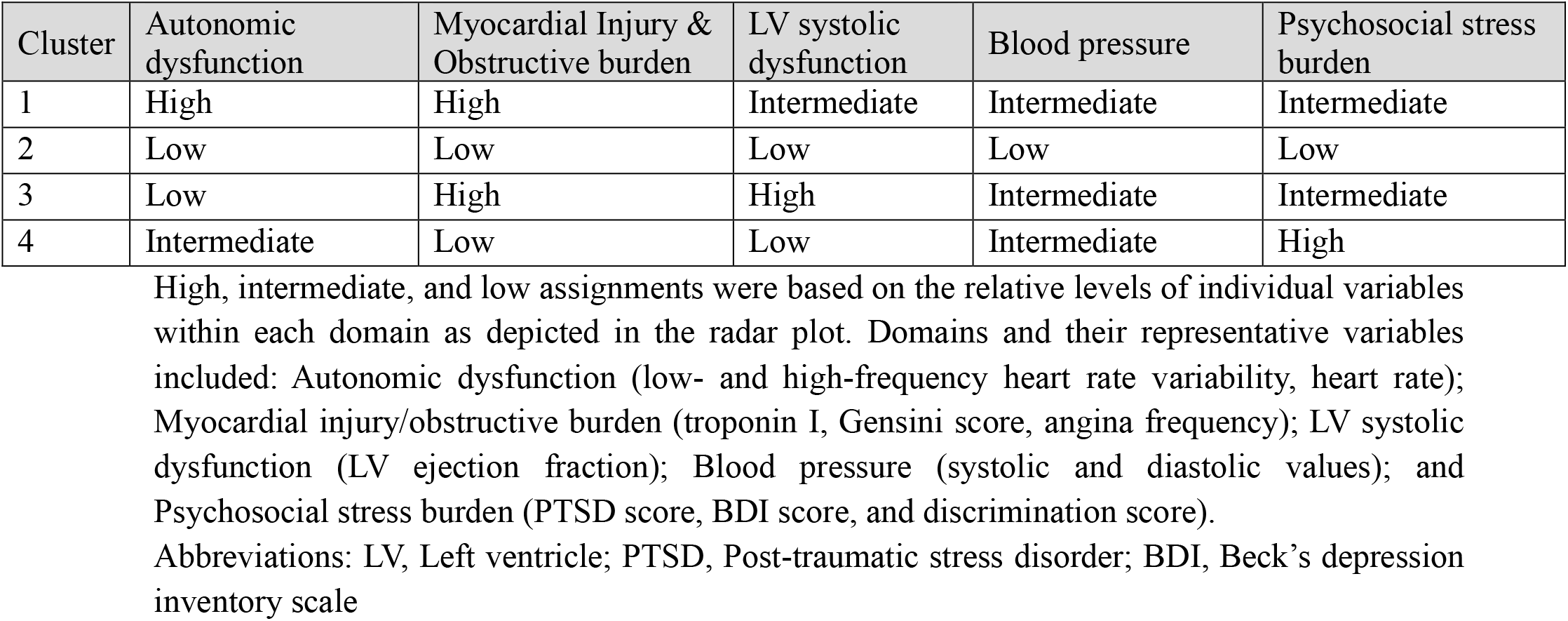
Summary of Phenotypic Domains for Each Phenogroup.

**Figure 2.**
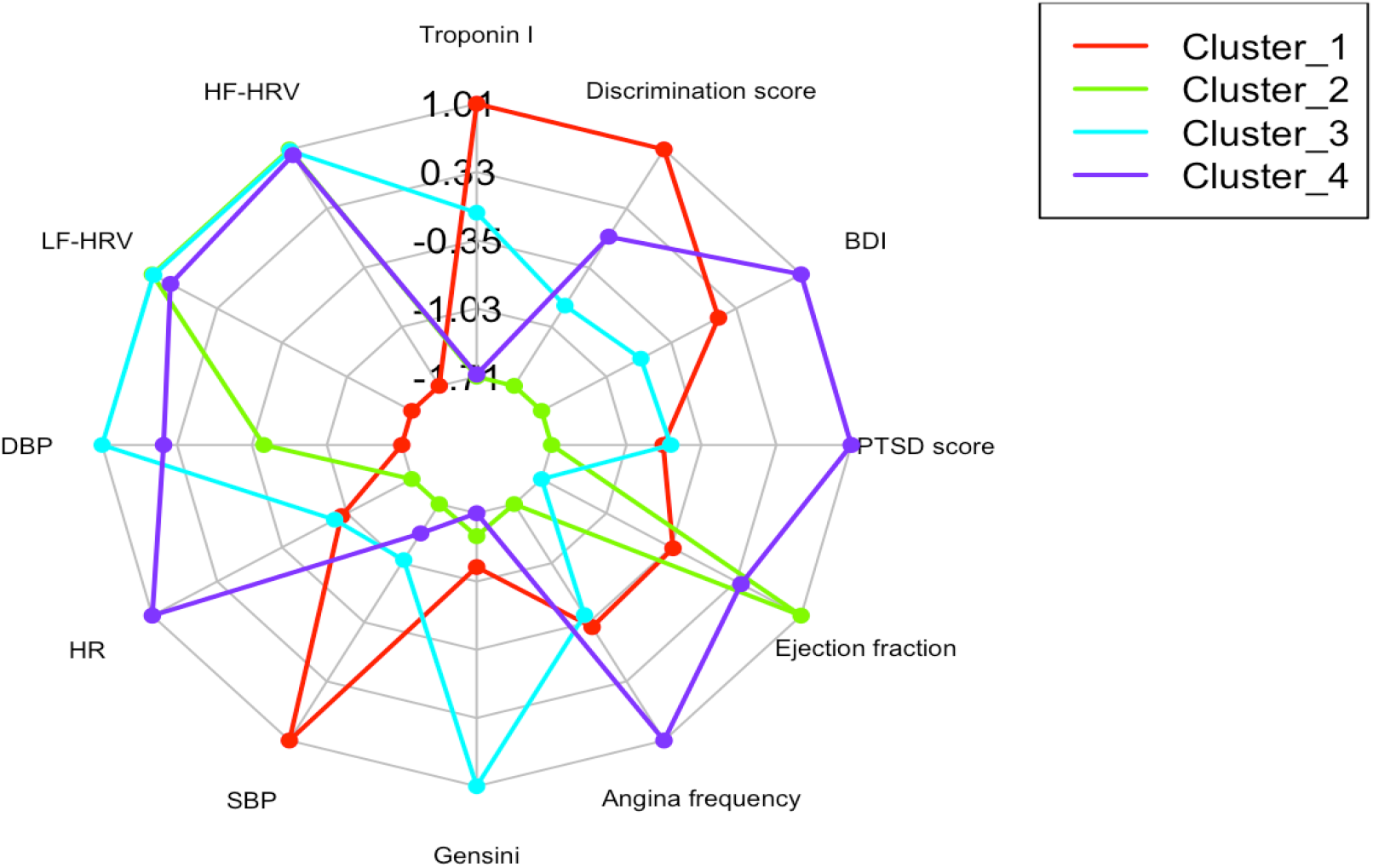
A Radar Chart showing the Pattern Variables in Each Phenogroup. Radar plot showing the mean values of variables for each cluster. Points closer to the center indicate lower values, while points farther from the center indicate higher values. Abbreviations: HF-HRV, High frequency heart rate variability; LF-HRV, Low frequency heart rate variability; DBP, Diastolic blood pressure; HR, Heart rate; SBP, Systolic blood pressure; BDI, Beck’s depression inventory score; PTSD, Post-traumatic stress disorder

### Clinical outcomes and prognostic significance

The cumulative incidences of CVD-specific and all-cause mortality were highest among participants in phenogroup 3 and lowest among those in phenogroup 2 (Figure 3). Using phenogroup 2 (“healthy-heart”) as the reference, markedly higher risks of both outcomes were observed in the other three groups across both crude and age-, sex-, and race-adjusted models. Participants in phenogroup 1 had a 3.7-fold higher risk of CVD-specific mortality and more than twofold increased risk of all-cause mortality compared with phenogroup 2 (Table 3). Phenogroup 3 carried the greatest risk overall, with nearly 8 to 10 fold higher risk (crude HR 9.7, 95% CI 3.4–28.1; adjusted HR 8.4, 95% CI 2.9–24.6) for CVD-specific mortality, as well as a 3.7-fold increased risk of all-cause mortality in both crude and adjusted models. Participants in phenogroup 4 had more than a twofold increased risk of CVD-specific mortality and a 40% higher risk of all-cause mortality, though not statistically significant (Table 3).

**Table 3.**
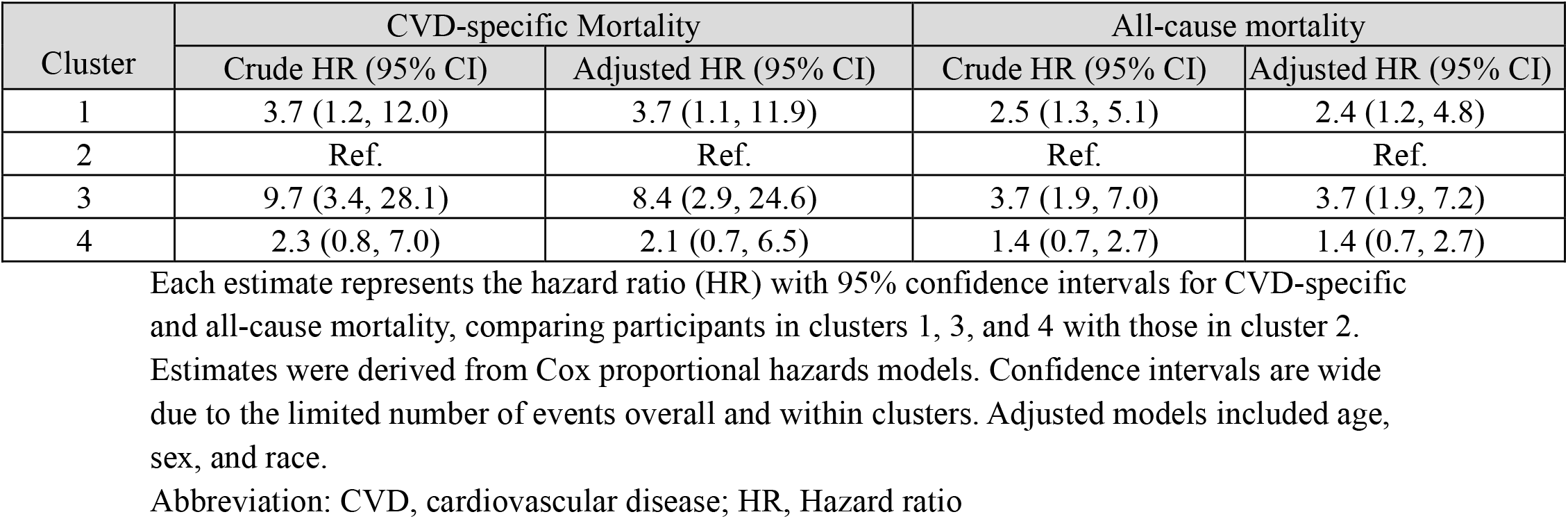
Association of Novel Phenogroups with CVD-specific and All-cause mortality.

| Cluster | CVD-specific Mortality |  | All-cause mortality |  |
| --- | --- | --- | --- | --- |
|  | Crude HR (95% CI) | Adjusted HR (95% CI) | Crude HR (95% CI) | Adjusted HR (95% CI) |
| 1 | 3.7 (1.2, 12.0) | 3.7 (1.1, 11.9) | 2.5 (1.3, 5.1) | 2.4 (1.2, 4.8) |
| 2 | Ref. | Ref. | Ref. | Ref. |
| 3 | 9.7 (3.4, 28.1) | 8.4 (2.9, 24.6) | 3.7 (1.9, 7.0) | 3.7 (1.9, 7.2) |
| 4 | 2.3 (0.8, 7.0) | 2.1 (0.7, 6.5) | 1.4 (0.7, 2.7) | 1.4 (0.7, 2.7) |

Each estimate represents the hazard ratio (HR) with 95% confidence intervals for CVD-specific and all-cause mortality, comparing participants in clusters 1, 3, and 4 with those in cluster 2.
Estimates were derived from Cox proportional hazards models. Confidence intervals are wide due to the limited number of events overall and within clusters. Adjusted models included age, sex, and race.
Abbreviation: CVD, cardiovascular disease; HR, Hazard ratio

**Figure 3.**
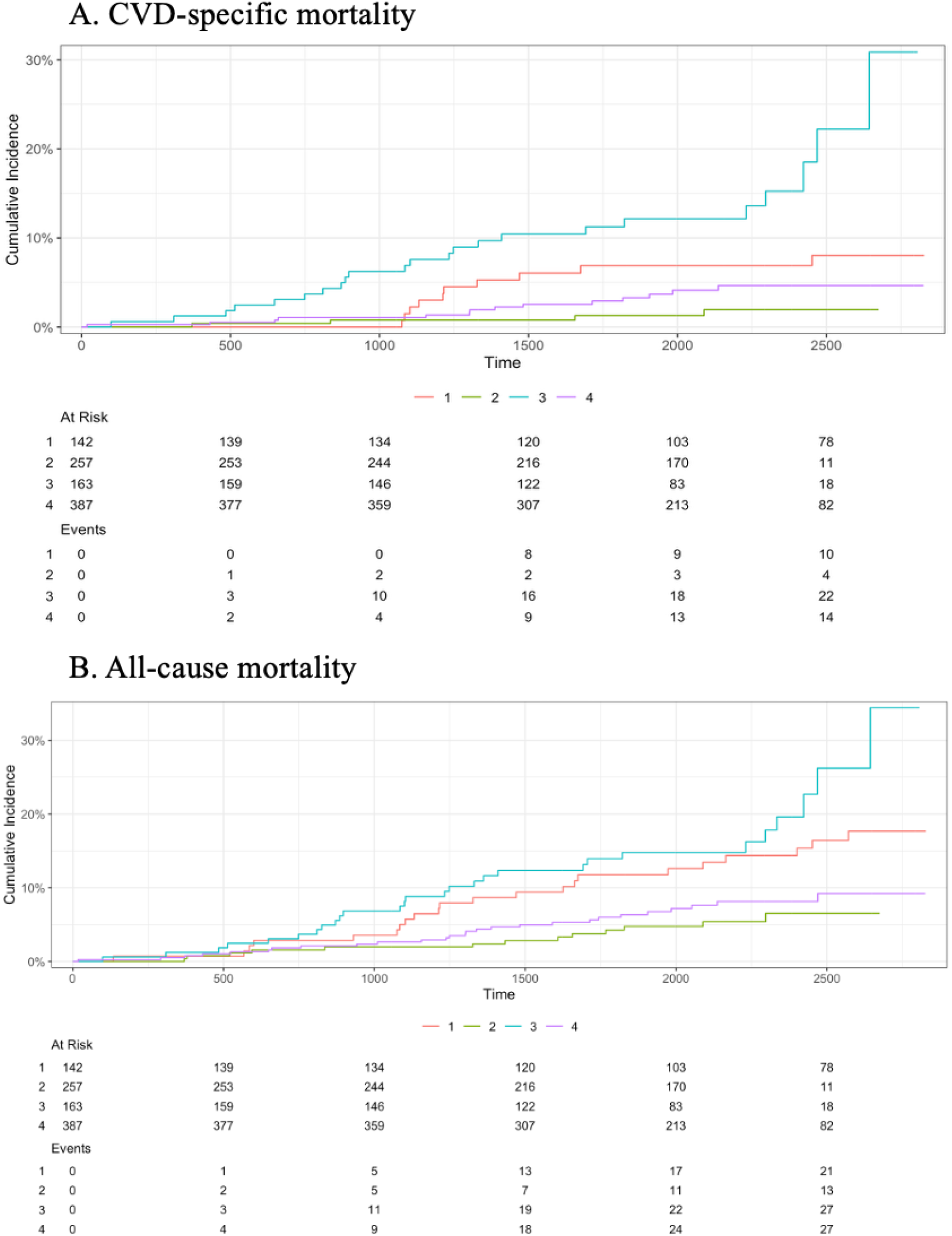
Cumulative Incidence curves for CVD-specific and all-cause Mortality by Phenogroups. Crude cumulative incidence curves for CVD-specific (A) and all-cause (B) mortality, stratified by the novel phenogroups.

## Discussion

In this study of over 900 participants with clinical CAD, we identified 4 primary phenogroups using model-based clustering which encompassed distinct pathophysiological patterns with different thematic elements, including, in increasing order of mortality risk: low-risk factor burden, high psychosocial burden, cardiac autonomic dysfunction and injury, and ischemic cardiomyopathy. To our knowledge, this is the first study to phenogroup patients with CAD with psychosocial and autonomic data, and our finding that both factors played a major role in group formation has important clinical implications as we consider individualized behavioral therapies.^20^ Although for the psychosocial group the association with CVD mortality was not statistically significant, the event rate was over two-folder higher than the low-risk group, and more research is warranted to better understand their prognosis and mechanisms of risk.

Previous research has shown that psychosocial stressors and autonomic dysfunction are predictive of outcomes,^5, 21^ but unfortunately such patients have been difficult to identify in clinical practice without rigorous testing. By shifting the paradigm to phenogroups, we can better identify individuals within these groups even when only partial data are available. For example, the psychological stress group also has a disproportionately high angina burden, yet low atherosclerotic burden. When treating angina in cases not explained by atherosclerosis, providers should also consider depression screening and biobehavioral therapies.^22^ Formal HRV testing is also not always available, but those in the dysautonomia group may be more easily identified when considering comorbidities that can affect autonomic function such as ischemic heart disease and diabetes. This group also had high perceived discrimination and moderate depressive symptoms, which are known contributors to dysautonomia.^23^

The “ischemic cardiomyopathy” phenogroup is the most concerning in terms of mortality risk, although in many clinical settings their high-risk status is already known. Nonetheless, many undiagnosed individuals in this group with low symptom burden may exist.^24^ This can occur especially in situations of significant barriers to care such as cost or distance from clinical settings. Other considerations include challenges of implementation of medical and lifestyle therapy what are already indicated by the guidelines. For example, a disproportionately low proportion of heart failure patients receive cardiac rehabilitation.^25^ Although this group did not have as high of a psychosocial burden, on average, as other groups, these risk factors are still important in this group and may increase their mortality risk even further.^26^

Although all patients with CAD should receive guideline directed medical therapy, their management from a health behavioral perspective calls for further discussion. For example, although cardiac rehabilitation is a guideline-based recommended therapy for certain patients with CAD, many individuals with stable ischemic forms do not qualify based on Medicare criteria.^27^ Depression screening is not universally recommended, and certain pharmacological treatments are controversial in this group. Biobehavioral treatments may be considered, but more implementation and outcomes research are needed to enable their mainstream use in clinical care. Additionally, neuromodulation therapies such as vagal nerve stimulation are potentially useful in the group with dysautonomia, but few studies have been carried out in patients with CAD.^28^

This study is subject to several limitations. The sample was drawn from a single urban metropolitan area, potentially limiting generalizability across certain ethnic and socioeconomic populations such as rural populations. External validation in independent cohorts is essential to confirm phenotype stability and clinical utility. Missing data was handled through multiple imputations, a technique which assumes missing-at-random mechanisms. Future studies should prospectively collect complete phenotypic data to avoid potential biases from missing data assumptions. Translation of phenotyping approaches into clinical practice requires development of automated tools that can rapidly compute phenotype probabilities from routine clinical data; thus far, not all metrics considered in this study are readily available. Nonetheless, such data are increasingly available, as seen in modern wristband wearable devices, for example.

## Conclusions

We were able to identify four distinct phenogroups amongst a group of stable coronary artery disease patients that integrate cardiac autonomic function, myocardial injury markers, hemodynamic parameters, and psychosocial factors into clinically meaningful groups. These findings may help facilitate individualized care strategies, especially for behavioral interventions. These findings support a more holistic model that considers psychosocial and behavioral underpinnings to CVD pathogenesis. This model may have significant impact on risk if the right therapies are prescribed, warranting more research to both validate these findings and apply them in clinical trials.

## Data Availability

All data produced in the present study are available upon reasonable request to the authors.

## Acknowledgements

We would like to thank the EPICORE, ECCRI, Emory cardiology, and Rollins School of Public Health staff for their tireless contributions to these studies.

## Sources of Funding

We would like to acknowledge the following funding sources: NIH/NHLBI K23 HL127251, R01 HL155711 for A.S., NIH/NHLBI R01 HL163998 and NIH/NHLBI R01 HL109413 for V.V., and P01 HL101398 for A.Q.

## Disclosures

The authors have no financial conflicts of interest to report.

## Notes

### Competing Interest Statement

The authors have declared no competing interest.

### Funding Statement

The study was funded NIH/NHLBI K23 HL127251, R01 HL155711 for A.S., NIH/NHLBI R01 HL163998 and NIH/NHLBI R01 HL109413 for V.V., and P01 HL101398 for A.Q.

### Author Declarations

IRB of Emory University gave ethical approval for this work

